# Validation of Serum Neurofilaments as Prognostic & Potential Pharmacodynamic Biomarkers for ALS

**DOI:** 10.1101/19002998

**Authors:** Michael Benatar, Lanyu Zhang, Lily Wang, Volkan Granit, Jeffrey Statland, Richard Barohn, Andrea Swenson, John Ravits, Carlayne Jackson, Ted M Burns, Jaya Trivedi, Erik P Pioro, James Caress, Jonathan Katz, Jacob L McCauley, Rosa Rademakers, Andrea Malaspina, Lyle W Ostrow, Joanne Wuu, on behalf of the CReATe Consortium

## Abstract

**Objective:** Identify preferred neurofilament assays, and clinically validate serum NfL and pNfH as prognostic and potential pharmacodynamic biomarkers relevant to ALS therapy development.

**Methods:** Prospective, multi-center, longitudinal observational study of patients with ALS (n=229), primary lateral sclerosis (PLS, n=20) and progressive muscular atrophy (PMA, n=11). Biological specimens were collected, processed and stored according to strict standard operating procedures (SOPs) ^1^. Neurofilament assays were performed in a blinded manner by independent contract research organizations (CROs).

**Results:** For serum NfL and pNfH measured using the Simoa assay, missing data (i.e. both technical replicates below the lower limit of detection (LLD) was not encountered. For the Iron Horse and Euroimmun pNfH assays, such missingness was encountered in ∼4% and ∼10% of serum samples respectively. Mean coefficients of variation (CVs) for pNfH in serum and CSF were ∼4-5% and ∼2-3% respectively in all assays. Baseline NfL concentration, but not pNfH, predicted the future ALSFRS-R slope and survival.

Incorporation of baseline serum NfL into mixed effects models of ALSFRS-R slopes yields an estimated sample size saving of ∼8%. Depending on the method used to estimate effect size, use of serum NfL (and perhaps pNfH) as pharmacodynamic biomarkers, instead of the ALSFRS-R slope, yields significantly larger sample size savings.

**Conclusions:** Serum NfL may be considered a clinically validated prognostic biomarker for ALS. Serum NfL (and perhaps pNfH), quantified using the Simoa assay, have potential utility as pharmacodynamic biomarkers of treatment effect.

## Introduction

Therapy development for amyotrophic lateral sclerosis (ALS) is challenging for many reasons, with the design and interpretation of phase II studies especially so ^2^. These “mid-development” studies typically employ traditional clinical measures such as survival or the rate of decline of the Revised ALS Functional Rating Scale (ALSFRS-R) as the principal measures of therapeutic effect, but are typically underpowered, making it difficult to decide which experimental therapeutics to advance from phase II to phase III ^3^. Biomarkers with prognostic and potential pharmacodynamic utility have great potential to help overcome these challenges ^3, 4^. Controlling for prognostic biomarkers, for example, might reduce phenotypic heterogeneity, thereby improving statistical power for a fixed sample size. Similarly, pharmacodynamic biomarkers may help to verify target engagement or demonstrate the presence of the intended biological effect. Neurofilaments – both light (NfL) and phosphorylated heavy (pNfH), in blood and cerebrospinal fluid (CSF) – have been proposed as potential ALS biomarkers with diagnostic value^5-9^, utility in predicting prognosis^6-8, 10-15^, and as possible pharmacodynamic biomarkers^16, 17^. The potential diagnostic utility aside, which is not the focus of this paper, the precise role of neurofilaments as prognostic or pharmacodynamic biomarkers in ALS trials is not yet fully defined. Several questions remain unresolved: Which pNfH assay, among the many available, should be selected? Do NfL and pNfH convey the same information, or should both be measured? Moreover, understanding the prognostic value of neurofilaments has been hampered by the use of *estimated* rather than *measured* rates of disease progression through longitudinal follow-up; and inconsistent consideration of the value added by neurofilaments to clinical predictors of prognosis (age, gender, bulbar onset, etc.). Similarly, studies of the longitudinal trajectories of neurofilaments have reached inconsistent conclusions about their relationship to clinical measures of disease progression, driven in part by relatively small participant numbers.

Here we present NfL and pNfH data (obtained from serum and CSF) from a large cohort of patients with ALS and related disorders who have undergone careful longitudinal clinical phenotyping along with serial collection of biological samples. Serum and CSF neurofilaments have been quantified using a variety of assays, performed by CROs blinded to participant identity and status. Combined with carefully collected phenotypic data, we have endeavored to address unanswered questions related to the prognostic and potential pharmacodynamic utility of neurofilament levels.

## Materials and Methods

### Study Population

Patients with ALS and related disorders were enrolled at multiple centers, through the *Phenotype-Genotype-Biomarker* study of the <u>C</u>linical <u>Re</u>search in <u>A</u>LS and Related Disorders for <u>T</u>h<u>e</u>rapeutic Development (CReATe) Consortium. Rigorously standardized clinical assessments and biological sample collections were performed every 3-6 months. Samples included in this experiment were from study visits that took place between April 2015 and November 2017.

### Standard Protocol Approvals, Registrations, and Patient Consents

The *Phenotype-Genotype-Biomarker* study is registered on clinicaltrials.gov (NCT02327845). The University of Miami IRB (the central-IRB for CReATe) approved the study; all participants provided written informed consent.

### Sample Collection, Processing and Storage

Blood was collected in serum-separating BD-vacutainers and allowed to clot upright at room temperature for 1-2 hours. Following centrifugation (1750g for 10 minutes at 4°C) serum was aliquoted into cryogenic sterile freestanding conical microtubes (Nalgene or Bio Plas Inc.) and stored at -80°C. CSF was collected in polypropylene tubes, centrifuged (1750g for 10 minutes at 4°C), aliquoted into polypropylene cryogenic sterile freestanding conical microtubes, frozen within ∼30 minutes of collection, and stored at - 80°C.

### Neurofilament Quantification

Serum and CSF neurofilament concentrations were quantified by Quanterix using their Simoa NfL^18, 19^ and pNfH assays^20^; each plate contained calibrators (0-500 pg/mL for NfL and 0-2000pg/ml for pNfH) and quality controls. Samples were diluted to fall within the range of the standard curve. Serum and CSF pNfH were also quantified by Iron Horse Diagnostics, using their in-house assay (which uses monoclonal capture and polyclonal detection antibodies)^11^ and the Euroimmun CE marked ELISA^21^ (which reverses the capture and detection antibodies). All samples were measured in duplicate at the same dilution, and all assays were performed blind to clinical parameters.

### Statistical Analysis

Symptom onset was defined as the first occurrence of limb weakness, dysarthria, dysphagia, or dyspnea. ΔFRS was calculated by subtracting baseline ALSFRS-R from 48 (maximum ALSFRS-R score), divided by time (months) from symptom onset to baseline ^22^. Survival duration was defined as time from symptom onset to PAV (≥22 hours/day non-invasive ventilation), tracheostomy or death.

Each sample was assayed twice to generate two replicates on each platform. We then computed (a) the number of samples for which neurofilament levels were below the LLD, (b) the mean difference and 95% confidence interval of the differences between replicates (i.e. limits of agreement ^23^), and (c) the coefficients of variation (CVs) between replicates. The association between serum and CSF concentrations was assessed using Spearman’s rank correlation. These analytic characteristics were qualitatively compared between assay platforms and guided our selection of one of the pNfH assays for further investigation.

To evaluate the prognostic utility of serum NfL and pNfH, we fitted a linear model to estimate the change in ALSFRS-R for each subject over time, using only the subset of participants with at least three ALSFRS-R scores. The associations between ALSFRS-R slope estimates and pre-specified clinical and baseline neurofilament predictor variables were investigated using multivariable regression. To assess the joint contribution of NfL and pNfH, a likelihood ratio test was used to compare the full model (including clinical and neurofilament variables) to a reduced model (including only clinical variables). The prognostic value of serum neurofilaments was also evaluated by Kaplan-Meier (univariate) and Cox regression (multivariate) methods, using PAV- and tracheostomy-free survival as the outcome.

We then estimated trajectories of changes in neurofilament levels over time using linear regression fitted separately to data from each subject. This model, which was limited to subjects with at least three available neurofilament values, included neurofilament as the outcome variable and time as predictor variable. Because neurofilament values were heavily skewed, natural logarithm-transformation was used to help achieve normality. We also estimated the impact of age in our cohort, using the same linear regression analysis described in Disanto et al. (2017). Briefly, a linear model with log neurofilament as the dependent variable, age as independent variable was fit to the baseline dataset. The estimated regression coefficient for age was then back transformed to the original scale, so that it reflects multiplicative effects (i.e., an estimate of 1.05 means an increase of ∼5% in neurofilament).

To assess the impact of considering baseline serum NfL on the sample size for a placebo-controlled clinical trial with ALSFRS-R slope as its primary outcome, we conducted a simulation similar to that described by Kuffner^24^. Briefly, we compared two mixed effects models, typically used for assessing treatment effects on ALSFRS-R slopes in clinical trials: Model-1 included ALSFRS-R scores as the outcome variable, time, time x group (treatment vs. controls), log(baseline NfL), time x log(baseline NfL) as fixed effects as well as random subject effects to account for correlations from repeated measures in the same subjects; Model-2 is the same as Model 1, but without variables involving NfL. 1000 simulations were performed. For each simulation, treatment group assignment was simulated at random, which assigned half of the patients with ≥3 ALSFRS-R scores (n=53) to the treatment group and the remaining to the control group. With *s*_1_ and s_2_ representing the estimated standard errors of time x group effects in Model-1 and Model-2, respectively, the percent reduction in sample size is proportional to the reduction in variance given by 100 * [1 - (*s*_1_ ^2^ /*s* _2_ ^2^)].

We also explored the utility of using serum NfL and pNfH as biomarkers to detect pharmacodynamic effect in a phase-2 clinical trial. Sample size estimations for such a trial required us to define a difference (or change) in serum neurofilament that might be regarded as clinically meaningful and which a future trial would need to be powered to detect. First, we considered the difference in serum neurofilament between fast and slow progressing patients (ALSFRS-R rate of decline >1 and <0.5 point/month, respectively), reasoning that this difference in neurofilament could be considered meaningful since it corresponds to a clinically important difference in rates of disease progression. We also considered an alternative approach, in which we defined a meaningful change based on a slope that is more negative (i.e. steeper decline) than the lower bound of the 95% confidence interval of neurofilament trajectories in untreated patients. All analyses were performed using SAS (version 9.4) and R software (https://www.r-project.org/).

### Data Sharing Statement

De-identified data will be shared immediately following publication.

## Results

### Study Population

A total of 260 patients (229 ALS, 11 PMA, and 20 PLS), with a total of 617 person-visits, were included in this study (Table 1). For evaluation of the pharmacodynamic potential of neurofilament measurements, analysis was restricted to the subset of 106 ALS, 3 PMA, and 4 PLS patients with at least 3 longitudinal biofluid collections (Table 1).

**Table 1.** Study Participant Characteristics

|  |  | Entire Study Cohort |  |  | Longitudinal Subset <sup>e</sup> |  |  |
| --- | --- | --- | --- | --- | --- | --- | --- |
|  | Descriptive Statistic | ALS (N=229) | PMA (N=11) | PLS (N=20) | ALS (N=106) | PMA (N=3) | PLS (N=4) |
| # of collections: | N (%) | -- | -- | -- |  |  |  |
| 3 collections |  |  |  |  | 53 (50%) | 2 (67%) | 1 (25%) |
| 4 collections |  |  |  |  | 34 (32%) | 0 | 2 (50%) |
| 5 collections |  |  |  |  | 19 (18%) | 1 (33%) | 1 (25%) |
| Baseline age (years) | Mean ± SD | 60.1 ± 11.8 | 56.8 ± 11.6 | 58.8 ± 11.1 | 59.6 ± 12.8 | 55.0 ± 7.5 | 56.3 ± 12.8 |
| Male | N (%) | 134 (59%) | 8 (73%) | 9 (45%) | 58 (55%) | 3 (100%) | 2 (50%) |
| Genotype: | N (%) |  |  |  |  |  |  |
| C9ORF72 |  | 27 (12%) | 0 | 0 | 9 ( 8%) | 0 | 0 |
| Other |  | 10 ( 4%) | 0 | 0 | 3 ( 3%) | 0 | 0 |
| Unknown |  | 192 (84%) | 11 (100%) | 20 (100%) | 94 (89%) | 3 (100%) | 4 (100%) |
| Site of onset: | N (%) |  |  |  |  |  |  |
| Bulbar only |  | 42 (18%) | 0 | 6 (30%) | 22 (21%) | 0 | 2 (50%) |
| Limb only |  | 151 (66%) | 11 (100%) | 9 (45%) | 71 (67%) | 3 (100%) | 2 (50%) |
| Other region(s) only |  | 4 ( 2%) | 0 | 0 | 2 ( 2%) | 0 | 0 |
| Multiple regions |  | 24 (11%) | 0 | 4 (20%) | 8 ( 7%) | 0 | 0 |
| Unknown |  | 8 ( 4%) | 0 | 1 ( 5%) | 3 ( 3%) | 0 | 0 |
| Onset to baseline (years) | Median (IQR) | 1.8 (1.0 – 3.2) | 2.4 (1.7 – 9.0) | 7.9 (3.8 – 11.6) | 1.9 (1.0, 3.9) | 2.4 (2.3, 3.9) | 5.5 (2.4, 10.5) |
| Diagnosis to baseline (years) | Median (IQR) | 0.7 (0.3 – 1.6) | 1.0 (0.6 – 4.3) | 2.5 (1.5 – 5.5) | 0.8 (0.4, 1.9) | 1.6 (1.3, 3.2) | 3.8 (1.2, 8.9) |
| Baseline ALSFRS-R | Mean (± SD) | 34.7 ± 7.5 | 33.5 ± 8.5 | 35.2 ± 8.1 | 35.4 ± 7.4 | 27.0 ± 13.0 | 33.8 ± 9.2 |
| Baseline ΔFRS | Mean (± SD) | 0.62 ± 0.48 | 0.34 ± 0.36 | 0.17 ± 0.12 | 0.52 ± 0.40 | 0.52 ± 0.42 | 0.25 ± 0.13 |
| Baseline serum NfL (pg/ml) <sup>a</sup> | Median (range) | 17 (2, 369) <sup>b</sup> | 7 (3, 26) | 10 (2, 54) | -- | -- | -- |
| Baseline serum pNfH (pg/ml) <sup>a</sup> | Median (range) | 67 (1, 976) <sup>b</sup> | 27 (2, 101) | 30 (1, 328) | -- | -- | -- |
| Baseline CSF NfL (pg/ml) <sup>a</sup> | Median (range) | 107 (12, 269) <sup>c</sup> | N/A | N/A | -- | -- | -- |
| Baseline CSF pNfH (pg/ml) <sup>a</sup> | Median (range) | 103 (16, 216) <sup>c</sup> | N/A | N/A | -- | -- | -- |
| Baseline to last Nf (months) | Median (IQR) | -- | -- | -- | 11.0 (7.0, 13.6) | 7.1 (6.9, 12.6) | 17.9 (11.5, 23.8) |
| ALSFRS-R slope | Mean (± SD) | -- | -- | -- | -0.65 ± 0.65 | -0.57 ± 0.14 | -0.09 ± 0.46 |
| Survival time (months) | Median (95%CI) | 25.9 (22.6, 31.4) | 24.6 (2.3, 33.7) | Not estimable <sup>d</sup> | 30.6 (25.7, 36.5) | 24.6 (2.3, 33.7) | Not estimable <sup>d</sup> |

<sup>a</sup> Neurofilament data generated through Simoa assays<sup>b</sup> Baseline serum data not available for 4 ALS participants<sup>c</sup> Baseline CSF data only available for 29 ALS participants<sup>d</sup> Median survival time could not be reliably estimated because only 2 PLS patients reached survival endpoint during study follow-up<sup>e</sup> Includes the subset of study participants with at least 3 longitudinal neurofilament measurements (and contemporaneously collected ALSFRS-R scores) available

### Analytic Characteristics of Neurofilament Assays in Serum and CSF

In CSF, both replicates were very infrequently below assay LLDs (i.e. undetectable); this was true for both NfL and pNfH and in all assays. For serum, undetectable values were infrequent using the Simoa NfL and pNfH assays, but were encountered in ∼4% and ∼10% respectively of samples assayed using the Iron Horse and Euroimmun pNfH platforms (Table 2). Agreement between technical replicates was high for all assays except for CSF analysis using the Euroimmun assay (Table 2). The mean CVs for NfL were ∼3% in serum and CSF; and for pNfH were ∼4-5% and ∼2-3% for all assays in serum and CSF, respectively. We selected the Simoa platform for subsequent analyses, given that it had the fewest undetectable values, and good replicate agreement (low CV) in both serum and CSF.

**Table 2.** Analytic Characteristics of Neurofilament Light (and Phosphorylated Neurofilament Heavy

|  | Neurofilament Light |  | Phosphorylated Neurofilament Heavy |  |  |  |  |  |
| --- | --- | --- | --- | --- | --- | --- | --- | --- |
|  | Simoa |  | Simoa |  | Iron Horse |  | Euroimmun |  |
|  | Serum | CSF | Serum | CSF | Serum | CSF | Serum | CSF |
| Total number of samples | 614 | 78 | 614 | 78 | 614 | 78 | 614 | 78 |
| Number of samples with value(s) below LLD: |  |  |  |  |  |  |  |  |
| In both replicates | 0 | 0 | 0 | 1 | 26 | 0 | 64 | 0 |
| In only one replicate | 4 | 1 | 15 | 2 | 36 | 0 | 73 | 0 |
| Difference (in pg/ml) between replicates, mean (95% CI) <sup>a</sup> | -0.05<br>(-4.0, 3.9) | -0.03<br>(-11.8, 11.7) | -0.4<br>(-26, 25) | 1.0<br>(-7, 9) | -0.8<br>(-16, 14) | -4.9<br>(-90, 80) | -0.02<br>(-38, 38) | 67.9<br>(-163, 298) |
| Coefficient of variation (CV) <sup>b</sup>, mean | 3.2 | 2.9 | 4.7 | 2.1 | 4.0 | 2.7 | 5.3 | 3.0 |
| Number (proportion) of samples with CV > 10 | 12<br>(2.0%) | 1<br>(1.3%) | 51<br>(8.3%) | 0<br>(0.0%) | 63<br>(10.3%) | 0<br>(0.0%) | 88<br>(14.3%) | 0<br>(0.0%) |

LLD = lower limit of detection
<sup>a</sup> Difference = replicate 2 – replicate 1. The 95% CI of this difference is also known as the limits of agreement.<sup>b</sup> CV = 100 x (standard deviation of replicates 1 and 2) / (mean of replicates 1 and 2)

### Baseline Neurofilament Concentrations

Initial serum concentrations of NfL were higher in the ALS group compared to those with PMA and PLS (p<0.001; Table 1). In the ALS group, higher serum NfL was associated with older age, higher ΔFRS, female gender, the presence of a *C9ORF72* repeat expansion, and bulbar symptom onset (all p<0.001), but not with baseline ALSFRS-R. Higher CSF NfL levels were similarly associated with older age, higher ΔFRS, and female gender, although not reaching statistical significance (possibly due to smaller sample size). Higher baseline serum pNfH among ALS patients was associated with older age (p=0.046), higher ΔFRS (p<0.001), female gender (p=0.002), and bulbar onset disease (p=0.035), but not *C9ORF72* repeat expansion status or baseline ALSFRS-R. Higher CSF pNfH levels were also associated with a higher ΔFRS, although not reaching statistical significance.

### Correlation between Serum and CSF Neurofilament Concentrations

While CSF could more directly reflect CNS pathophysiology, serum is more easily accessible. Therefore, we examined for each assay how well the value of serum measurements could serve as a proxy for CSF measurements. For pNfH, the serum-CSF correlations were comparable among the 3 assays albeit with weak correlations (r=0.16 to 0.22); the serum-CSF correlation was, however, much stronger for NfL (r=0.62, Simoa platform only).

### Potential Prognostic Utility of Baseline Neurofilament Concentrations

The average (±SD) rate of decline of the ALSFRS-R was 0.65 (±0.65) points per month. Multivariate regression analysis, with the ALSFRS-R slope as the outcome, considering potential clinical predictors of prognosis (age, gender, *C9ORF72* status, site of disease onset, baseline ALSFRS and ΔFRS) as well as baseline serum NfL and pNfH, identified only ΔFRS as a meaningful clinical predictor. For every 1-point increase in the ΔFRS (pre-slope), the ALSFRS-R rate of decline is worsened by an additional 0.43 points/month (p=0.006) (Table 3). The addition of baseline serum NfL to the model shows that this biomarker adds prognostic value that is not already explained by known clinical predictors. For every 1-point increase in log serum NfL level, the ALSFRS-R rate of decline is worsened by an additional 0.42 points/month (p<0.001). By contrast, baseline serum pNfH provided little additional prognostic value beyond the effects of clinical predictors (p=0.17). When both NfL and pNfH were jointly considered, the results showed that, together, these biomarkers added significantly more prognostic values independent of clinical predictors (p=0.002, likelihood ratio test); this result, however, was primarily driven by the effect of NfL.

**Table 3.** Impact of Clinical and Neurofilament Characteristics on ALSFRS-R Rate of Decline and Survival

| Covariates | Impact on ALSFRS-R Slope <sup>a</sup> |  | Impact on Survival <sup>b</sup> |  |
| --- | --- | --- | --- | --- |
|  | Estimate (95% CI) | p-value | Hazard Ratio (95% CI) | p-value |
| Baseline age (years) | 0.001<br>(-0.008, 0.010) | 0.89 | 1.02<br>(1.00, 1.05) | **0.049** |
| Male | -0.09<br>(-0.33, 0.15) | 0.47 | 1.37<br>(0.84, 2.23) | 0.21 |
| *C9ORF72* HREM | -0.16<br>(-0.57, 0.25) | 0.45 | 1.70<br>(0.91, 3.18) | 0.10 |
| Site of Onset: Bulbar | -0.04<br>(-0.45, 0.37) | 0.84 | 1.10<br>(0.53, 2.29) | 0.80 |
| Site of Onset: Limb | -0.01<br>(-0.46, 0.45) | 0.97 | 1.04<br>(0.48, 2.27) | 0.91 |
| Baseline ALSFRS-R | N/A | N/A | 0.96<br>(0.93, 0.99) | **0.02** |
| Baseline ΔFRS (points/month) | -0.43<br>(-0.73, -0.13) | **0.006** | 1.67<br>(1.09, 2.57) | **0.02** |
| Baseline log(NfL) (log(pg/ml)) | -0.42<br>(-0.62, -0.21) | **< 0.001** | 2.12<br>(1.39, 3.23) | **< 0.001** |
| Baseline log(pNfH) (log(pg/ml)) | 0.07<br>(-0.03, 0.17) | 0.17 | 1.01<br>(0.84, 1.22) | 0.88 |

CI = confidence interval. N/A = not applicable.
<sup>a</sup> Based on multivariate regression analysis, with the ALSFRS-R slope (i.e. ALSFRS-R rate of decline) as outcome
<sup>b</sup> Based on Cox proportional hazards model, with tracheostomy- and permanent assisted ventilation (PAV)-free survival as outcome

In univariate survival analyses, the presence of *C9ORF72* repeat expansion, higher ΔFRS at baseline, and higher baseline serum NfL levels, were each associated with worse prognosis (shorter survival), but baseline serum pNfH levels were not (Figure 1). In a multivariate Cox proportional hazards model that considered age, gender, *C9ORF72* status, site of disease onset, baseline ALSFRS-R, ΔFRS, and baseline serum levels of NfL and pNfH, only age, baseline ALSFRS-R, ΔFRS, and serum NfL were significant predictors of survival (Table 3).

**Figure 1.**
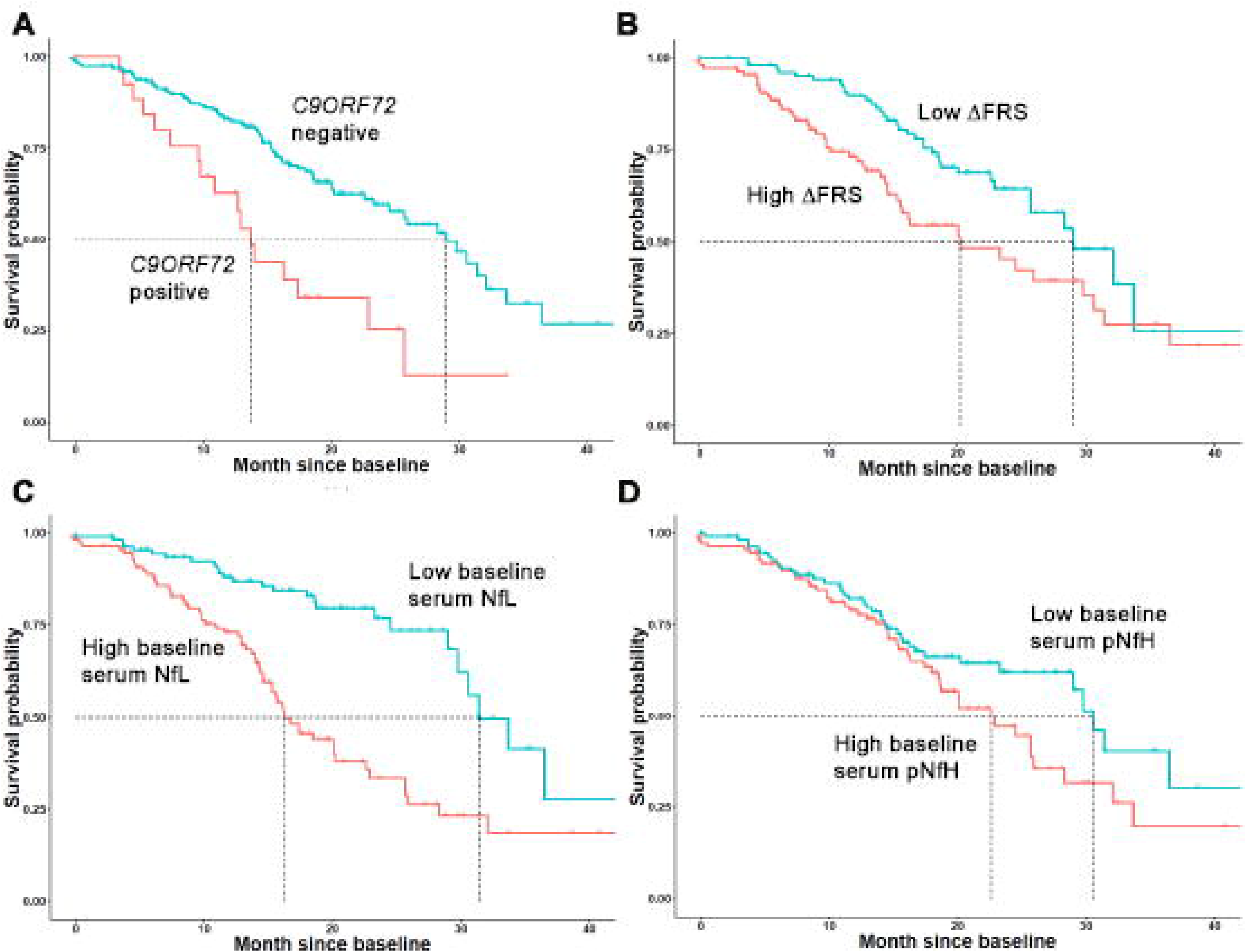
Kaplan-Meier survival curves, showing the prognostic value of (A) *C9ORF72* repeat expansion, (B) ΔFRS (dichotomized at the median, 0.62points/month), (C) baseline serum NfL (dichotomized at the median, 17pg/ml), and (D) baseline serum pNfH (dichotomized at the median, 67pg/ml). The presence of a *C9ORF72* repeat expansion, higher ΔFRS, higher baseline serum NfL, and higher baseline serum pNfH are shown in red.

### Potential Pharmacodynamic Utility of Longitudinal Neurofilament Concentrations

To ready the use of serum NfL and pNfH as potential pharmacodynamic biomarkers of treatment effect in future trials, we estimated their longitudinal trajectories (and variability of slopes). For serum NfL, the average slope was 0.011 log units/month (95% confidence intervals [CI]: -0.054, 0.076). Consistent with prior studies ^25^, we also observed a positive association between NfL and age, with 1.3% increase in NfL for each additional year (95% CI = 0.4%-2.3%). For serum pNfH, the average slope was 0.006 log units/month (95% CI: -0.063, 0.084) (Figure 2). We also quantified the magnitude change in biomarker concentration on the raw scale compared to baseline (Figure 3). For serum NfL, the 90^th^ and 95^th^ percentiles of the maximum absolute changes in concentration from baseline are 15 and 22pg/ml respectively. For serum pNfH, the 90^th^ and 95^th^ percentiles of the maximum absolute changes in concentration from baseline are 146 and 196pg/ml respectively.

**Figure 2.**
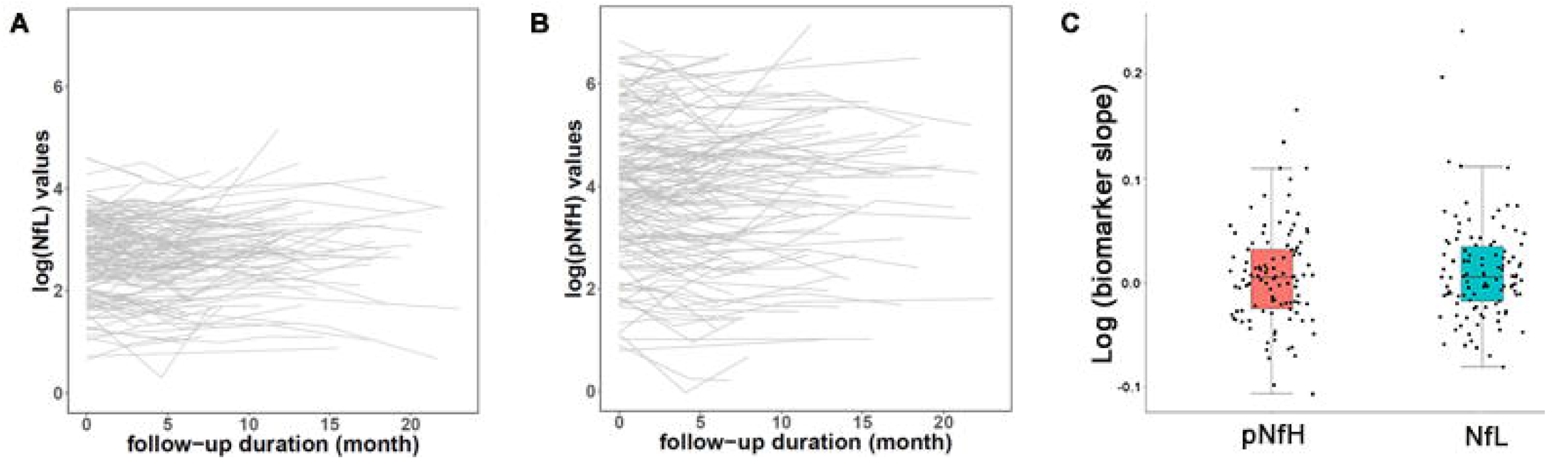
Longitudinal trajectories of serum neurofilaments: (A) spaghetti plot of log-transformed neurofilament light level (NfL), (B) spaghetti plot of log-transformed phosphorylated neurofilament heavy level (pNfH), and (C) boxplot of the distribution of NfL and pNfH slopes. Slope estimates were obtained from a linear model with log-transformed neurofilament level as the outcome, and time as the independent variable.

**Figure 3.**
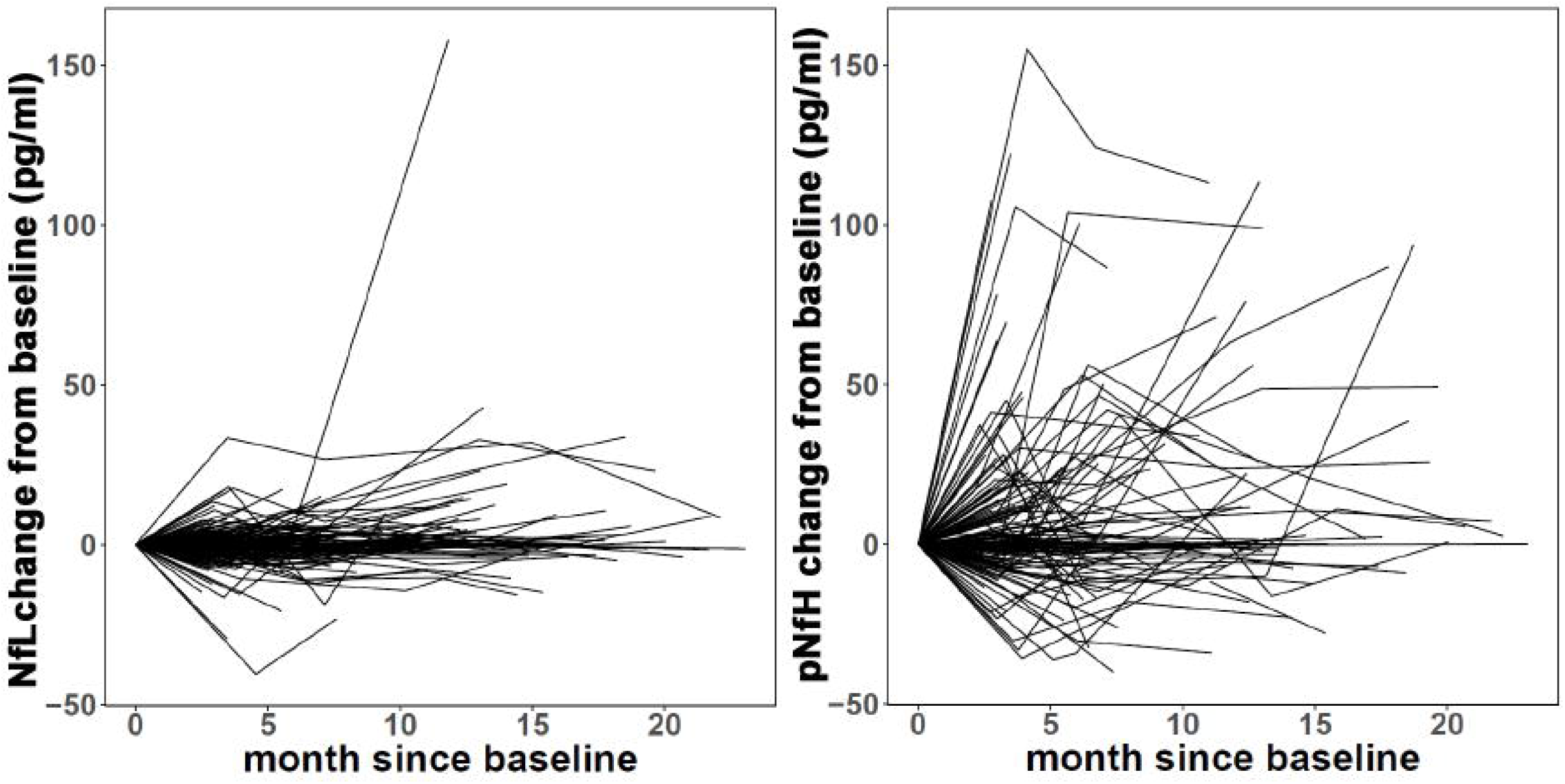
Change in serum neurofilaments over time, as compared to baseline level: (A) Change from baseline serum neurofilament light (pg/ml), and (B) change from baseline in serum phosphorylated neurofilament heavy (pg/ml).

### Utility of Neurofilament Biomarkers in Reducing Sample Size for Future Clinical Trials

Since baseline NfL level is predictive of ALSFRS-R trajectories, we hypothesized that the number of patients needed for a trial would be reduced when baseline NfL levels, as a *prognostic* biomarker, are considered. This may be done by including serum NfL as a covariate in the linear mixed models typically used to examine ALSFRS-R slopes. Using a simulation study similar to that described by Kuffner^24^, we found that the sample size for an ALS trial would be reduced by about 8.2% with the addition of baseline NfL measurements.

Furthermore, we estimated the sample size required for a future phase-2 trial with serum NfL (or serum pNfH) as a *pharmacodynamic* biomarker. We considered 2 different outcome measures and what a clinically meaningful treatment difference may be for each: First, assuming that an experimental therapeutic would need to change the *slope* of serum NfL from our observed average in the untreated state (0.011 log units/month) to less than the lower bound of the 95% confidence interval (−0.054 log units/month) and thereby an estimated treatment difference of 0.065 log units/month, with an estimated standard deviation of 0.048 log units/month, a total sample size of N=26 (13 treatment, 13 placebo) would provide 90% power to detect such a treatment difference, using a two-sample t-test with 5% significance level. Similarly, assuming that an experimental therapeutic would need to change the *slope* of serum pNfH from our observed average in the untreated state (0.006 log units/month) to less than the lower bound of the 95% confidence interval (−0.063 log units/month) and thereby an estimated treatment difference of 0.069 log units/month, with an estimated standard deviation of 0.046 log units/month, a total sample size of N=22 (11 treatment, 11 placebo) would provide 90% power to detect such a treatment difference, using a two-sample t-test with 5% significance level.

Alternatively, one may wish to achieve a clinically meaningful difference in NfL (or pNfH) *levels* (rather than trajectories). Based on an estimated treatment difference of 0.67 log(pg/ml)—which was our observed difference in baseline log(NfL) levels between fast progressors (ALSFRS-R decline of >1 point/month) and slow progressors (ALSFRS-R decline of <0.5 points/month), which were 3.10 and 2.43 log(pg/ml), respectively—and a standard deviation of ∼0.81 log(pg/ml), a total sample size of N=64 (32 treatment, 32 placebo) would provide 90% power to detect such a treatment difference, using a two sample t-test with 5% significance level. By contrast, the much smaller difference of 0.22 log(pg/ml) between serum pNfH levels among fast and slow progressors [3.82 log(pg/ml) and 3.60 log(pg/ml) respectively], combined with a standard deviation of ∼1.43 log(pg/ml), yields a total sample size of N=1778 (889 treatment, 889 placebo) in order to provide 90% power to detect such a treatment difference, using a two sample t-test with 5% significance level

For comparison, we also similarly estimated the required sample size for a clinically meaningful 20-30% reduction in *ALSFRS-R slope*, a common outcome measure employed in ALS clinical trials ^26^. Based on our observed average ALSFRS-R slope of - 0.648 points/month and an estimated standard deviation of 0.65 points/month, a total of N=1054 or N=470 patients would be needed respectively to provide power for detecting a 20% or 30% reduction in ALSFRS-R slopes.

## Discussion

While much has been written about the prognostic and potential pharmacodynamic utility of neurofilaments in ALS ^7, 12, 13, 21, 27^, most studies have been single-center, measured either NfL or pNfH (but not both), used only a single assay to quantify neurofilament levels, evaluated either blood or CSF (but not both), explored either survival or functional decline (but not both), and rarely quantified functional decline using prospectively collected ALSFRS-R data. Here, we have undertaken a multi-center study with head-to-head comparison of three different pNfH assays (performed by two different CROs) in serum and CSF, as well as an evaluation of the prognostic and potential pharmacodynamic utility of serum NfL and pNfH, using multivariate analytic techniques that explore the potential value neurofilaments add to readily available clinical information. Functional decline was quantified using prospectively collected ALSFRS-R data, as would be done in a clinical trial. We have also illustrated how serum neurofilament data might be used to aid the design and implementation of future phase II clinical trials.

For pNfH, the Simoa assay had the best sensitivity (fewest values below the assay LLD) and reproducibility (lowest CV between technical replicates in both serum and CSF). Using the Simoa assay, we also found a much stronger correlation between serum and CSF for NfL than for pNfH. The correlations between serum and CSF in this study are substantially lower than prior published reports ^12, 21, 28^. One possible explanation for this discrepancy is that we restricted our analyses to baseline data. As discussed by Bland and Altman, when multiple measurements taken from the same subjects are used to compute correlation coefficients, the result can appear artificially high because variability between measurements on the same subject are not properly accounted for and the independence assumption of correlation is violated ^23^.

We also found that serum NfL is prognostic of future ALSFRS-R decline and survival duration, providing information that is not captured by readily available clinical predictors. While absolute values can vary between patients, serum NfL levels remain largely stable in each subject over time. This may portend clinical utility as pharmacodynamic biomarkers if there are detectable changes in levels following exposure to an experimental therapeutic. This expectation is supported by recent studies of Nusinersen for spinal muscular atrophy (SMA) ^29, 30^, though definitive proof in adult motor neuron diseases would necessarily require an effective therapeutic to test. Moreover, we have illustrated the ways in which use of serum NfL in a phase-2 trial might permit a reduction in sample size. Incorporation of serum NfL as a covariate in a trial that uses the ALSFRS-R rate of decline as the primary outcome measure would yield a modest benefit (∼8% reduction in sample size), but use of NfL as a pharmacodynamic biomarker might offer meaningful sample size savings compared to more traditional phase-2 studies in which changes in the ALSFRS-R are used as the primary outcome measure. The potential utility of serum pNfH is more nuanced. On the one hand, we have found that it does not add prognostic value to readily available clinical parameters and serum NfL. On the other hand, the stability of serum pNfH suggests potential value as a pharmacodynamic biomarker but, depending on the approach used to quantify the clinical meaningfulness of a reduction in pNfH, this may or may not yield sample size savings in a phase-2 clinical trial setting.

The main limitation of this study is that the population may not be adequately representative of ALS patients who would ordinarily be enrolled in a clinical trial. Patients included in the current study were skewed towards those with more slowly progressive disease (evidenced by an ALSFRS-R slope of -0.65 points/month). Achieving better representation of a trial-like population is the goal of an ongoing study. We note, however, that the longitudinal stability of neurofilaments is not a consequence of this characteristic of the patient cohort, but rather a function of the relatively long latency from symptom onset to study enrollment and baseline assessment, which in turn reflects the well-described diagnostic delay that is characteristic of ALS ^31^. A fuller picture temporal course of the rise in neurofilament levels has emerged from studies of people at genetic risk for ALS, which have shown that NfL and pNfH rise in both the pre-symptomatic and early symptomatic periods ^32, 33^, with an expected plateau by the time of enrollment in a clinical trial or cohort study such as the one described herein.

A second limitation of the current study is that we have insufficient data to reliably comment on the value of CSF NfL and pNfH as either prognostic or pharmacodynamic biomarkers. Notwithstanding these considerations, this was a large multi-center study with prospective, systematic follow-up and strict SOPs for sample collection, processing and storage. All assays were performed by independent CROs blinded to clinical data, with comparative data generated across assay platforms, CROs and neurofilament types.

These results support two conclusions. First, serum NfL, but not serum pNfH, may be considered a clinically validated prognostic biomarker. Second, serum NfL (and perhaps pNfH) may have value as potential pharmacodynamic biomarkers, and both should be incorporated into ongoing and future phase-2 trials; the actual pharmacodynamic utility of these biomarkers, however, will only become apparent once we have truly effective treatments for ALS.

## Data Availability

De-identified data will be shared immediately following publication.

## Appendix 1 Authors

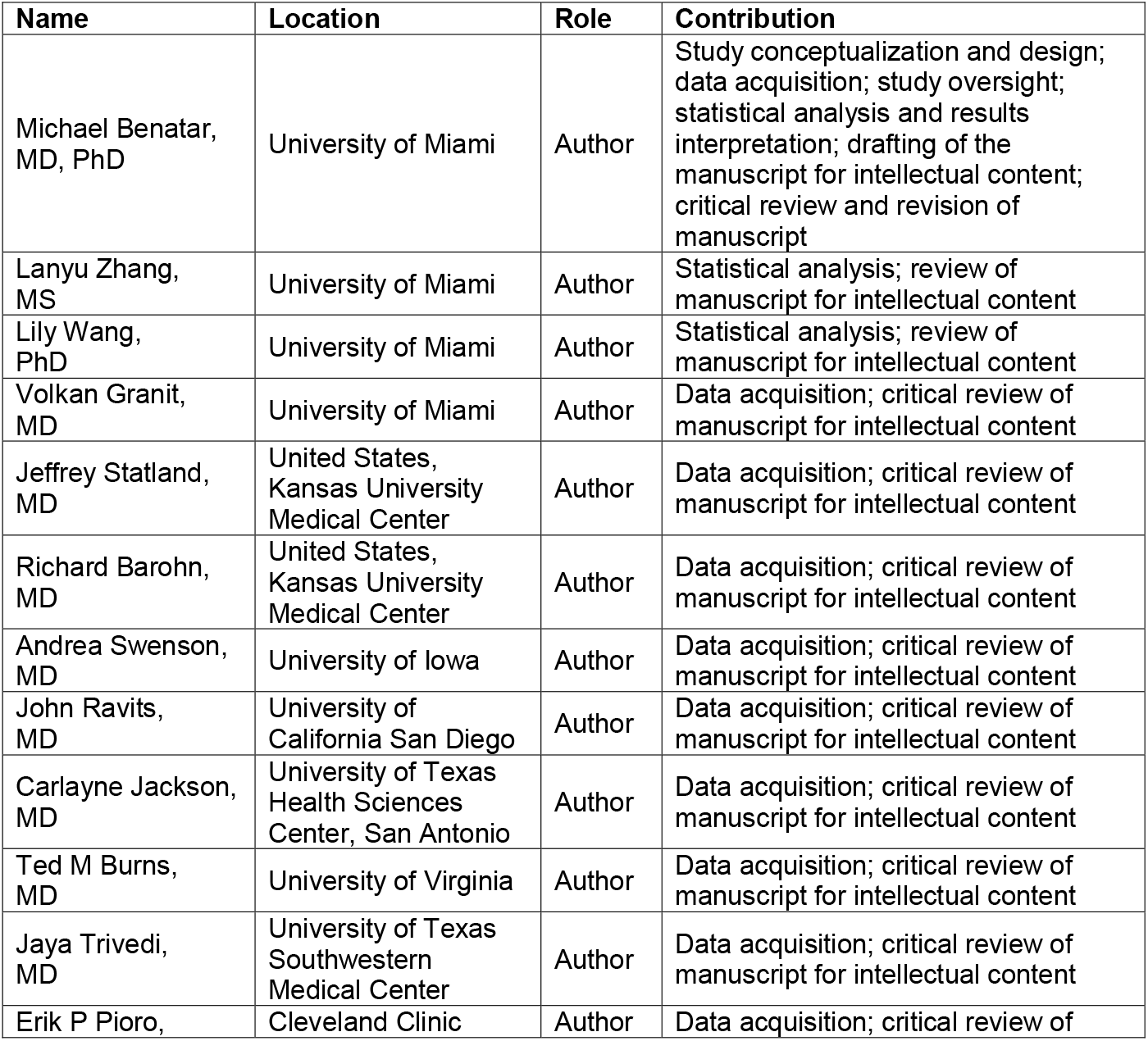
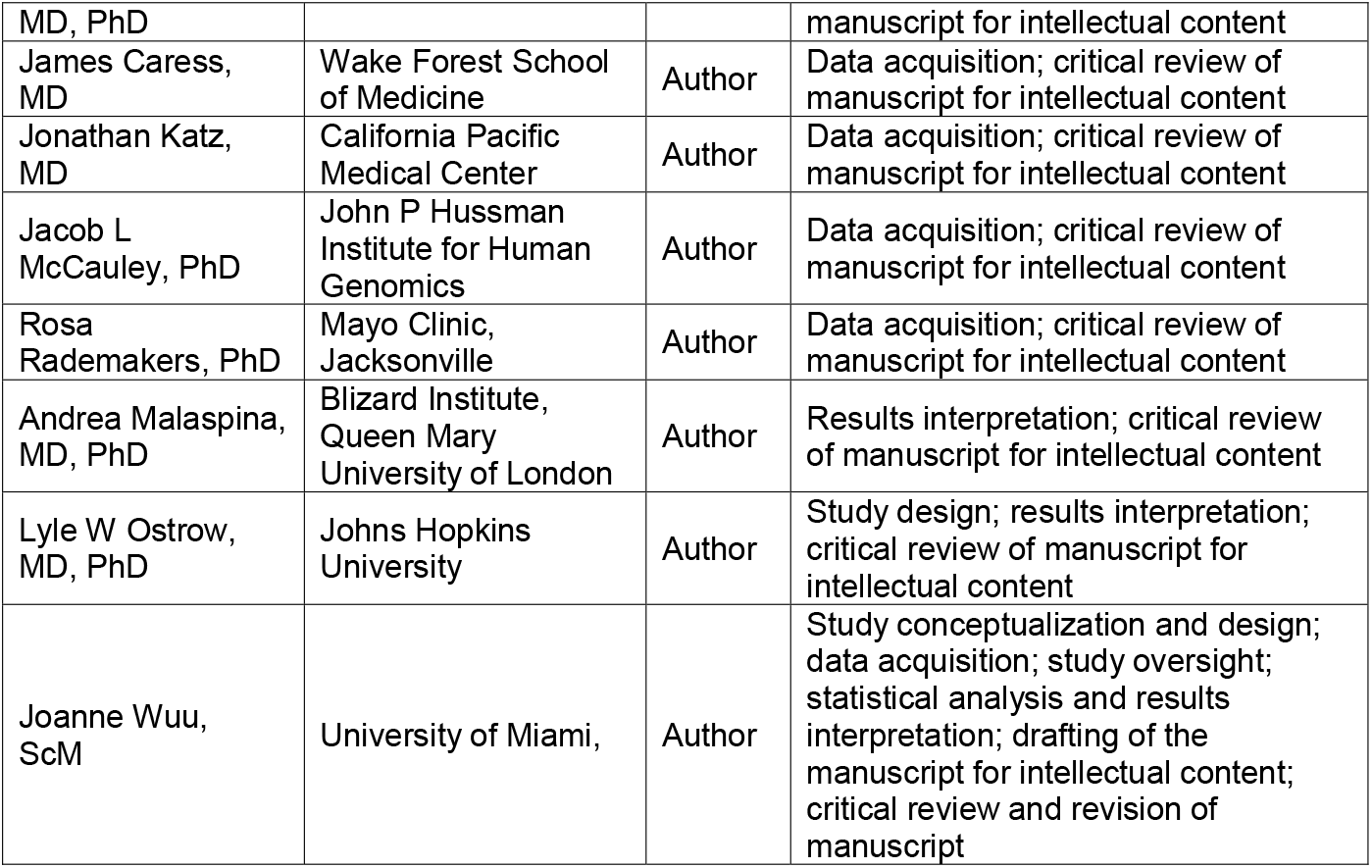

## Appendix 2 Co-Investigators (CReATe Consortium)

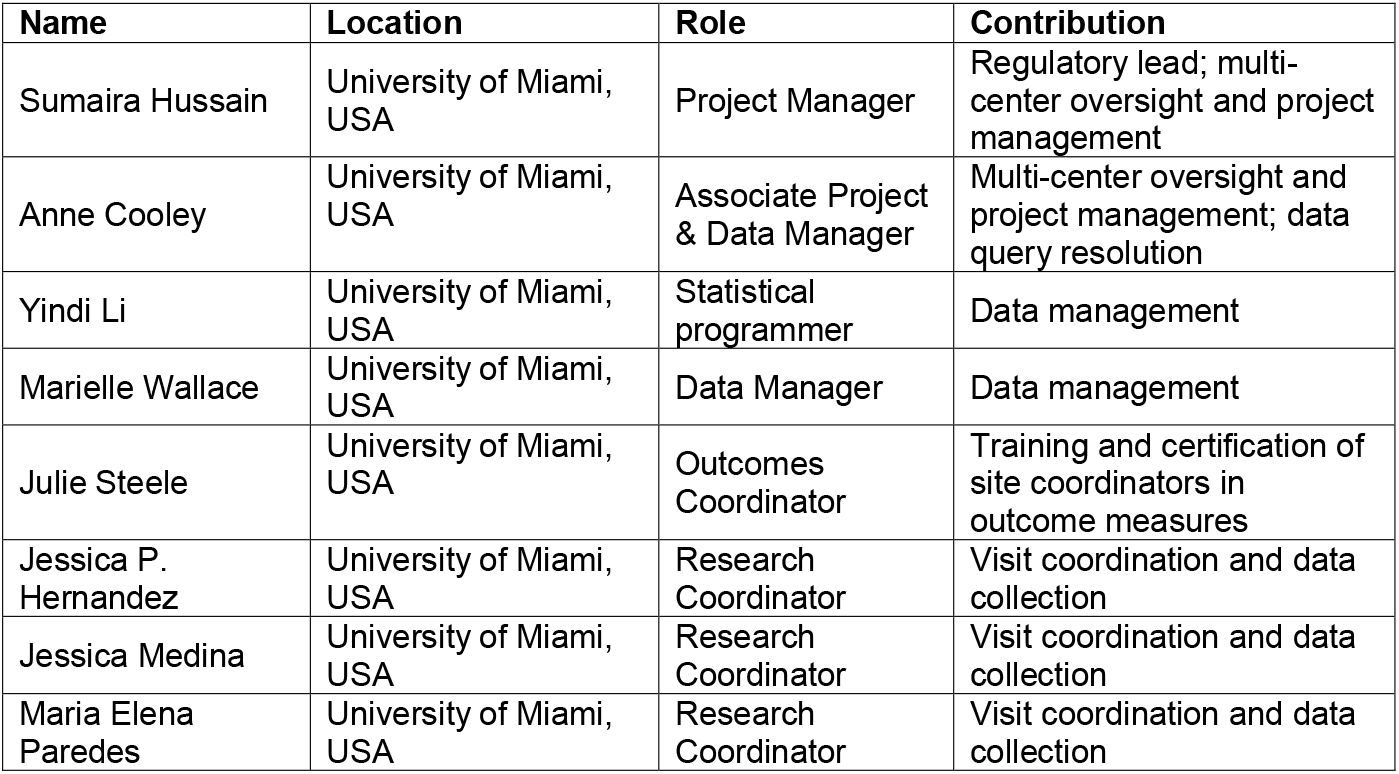
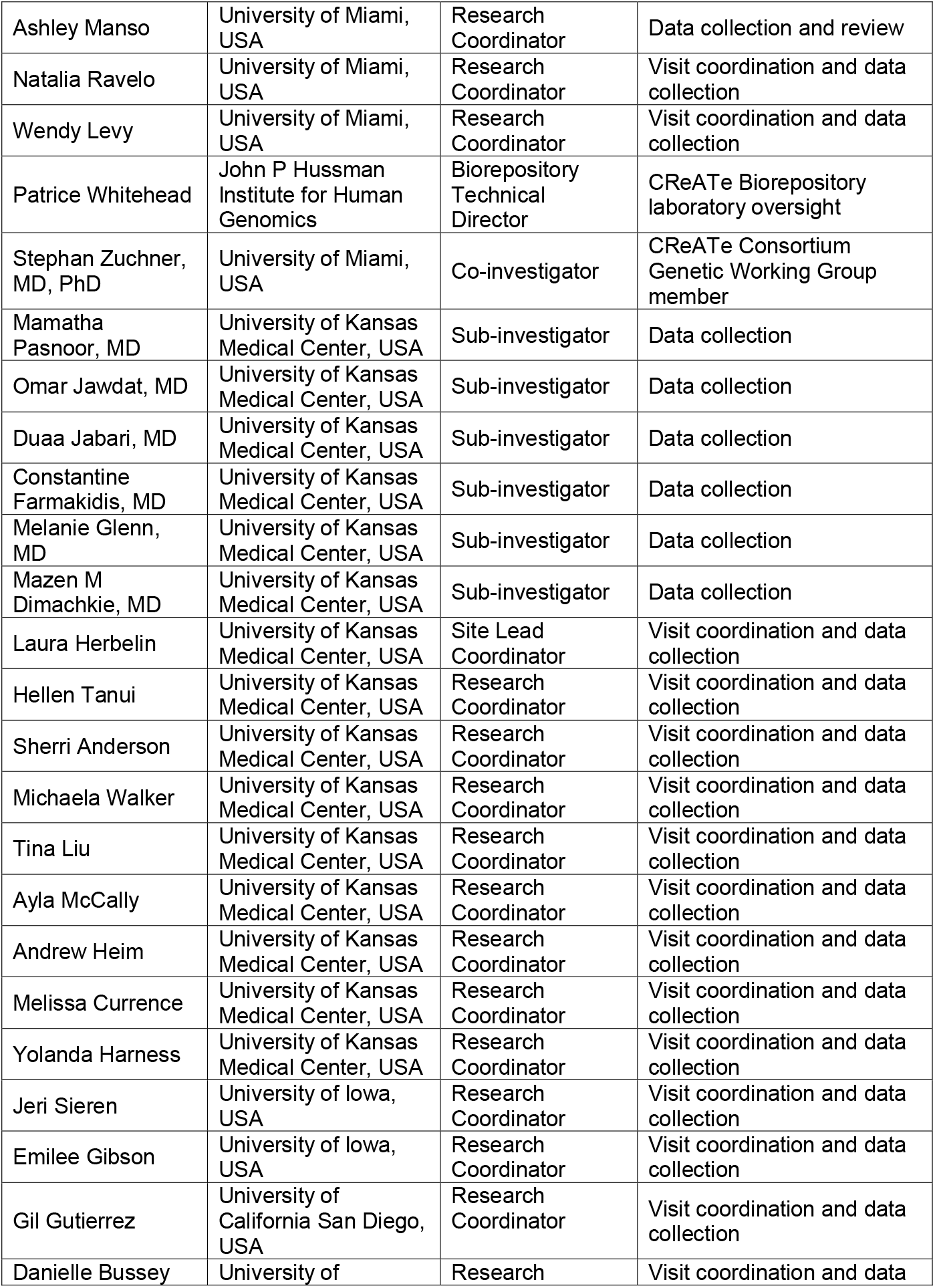
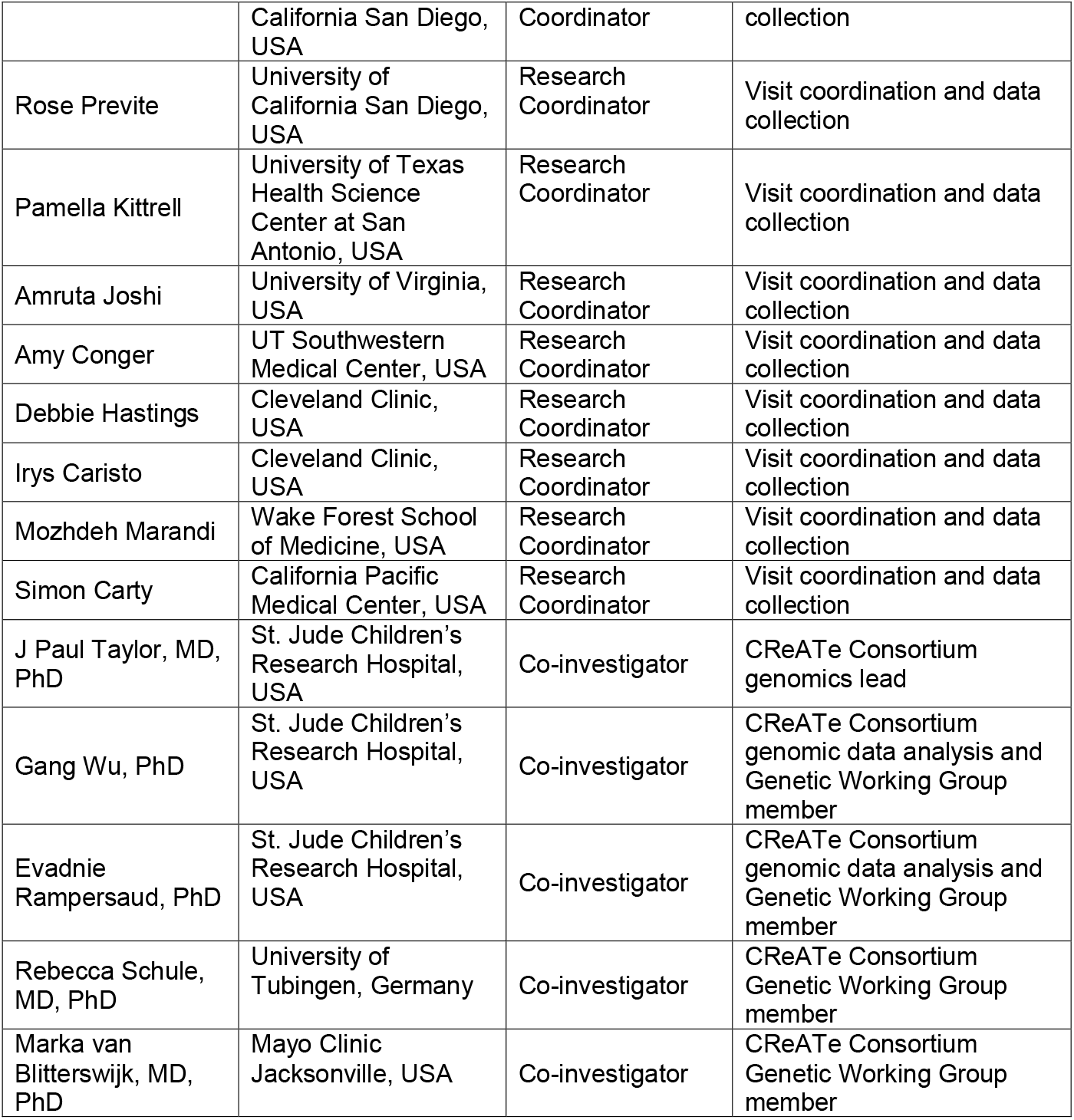

## Acknowledgements

We would like to acknowledge Joaquin del Cueto (Project Manager for Research Support, University of Miami); Dr. Lucie Bruijn (ALS Association Chief Scientist); Drs. Amelie Gubitz and Robin Conwit (NINDS Program Directors); Dr. Tiina Urv (NCATS Program Officer); Dr. Manish Raisinghani (CEO, Target ALS Foundation); and the Rare Diseases Clinical Research Network (RDCRN) Data Management and Coordinating Center (DMCC) at University of South Florida (PI: Dr. Jeffrey Krischer; regulatory specialists: Julie Martin and Kaleena Dezsi; and project managers: Callyn Kirk, Devon Rizzo, and Kristina Bowles). We would also like to thank Drs. Zaven Kaprielian (Amgen), Danielle Graham and Toby Ferguson (Biogen), Joe Lewcock (Denali), Felix Yeh (Genentech), Sophie Batteur Parmentier (Merck), Bryan Hill (Mitsubishi Tanabe Pharma), Dave Frendewey (Regeneron), Aarti Sharma (Regeneron), James Dodge (Sanofi), Laure Rosen (Takeda), Ian Reynolds and Neta Zach (Teva), Kelly Dakin (FBRI), Nadine Tatton (AFTD), Amanda Haidet-Phillips (MDA), Robert Bowser (Barrow Neurological Institute) and Leonard Petrucelli (Mayo Clinic Jacksonville) for helpful discussions around selecting biomarker candidates and contract research organizations. We would also like to acknowledge the staff and support of the John P. Hussman Institute for Human Genomics Biorepository Facility at the University of Miami School of Medicine. Most importantly, we are indebted to patients who participated in the CReATe Phenotype-Genotype-Biomarker (PGB) study for their altruism as well as their commitment and contribution to advancing therapeutic development for ALS and related disorders.

